# Endovascular Thrombectomy versus Intravenous Alteplase for Distal Vessel Occlusions: A Propensity Score-Matched Analysis

**DOI:** 10.1101/2023.10.22.23297380

**Authors:** Tomohide Yoshie, Toshihiro Ueda, Yasuhiro Hasegawa, Masataka Takeuchi, Masafumi Morimoto, Yoshifumi Tsuboi, Ryoo Yamamoto, Shogo Kaku, Ayabe Junichi, Takekazu Akiyama, Daisuke Yamamoto, Kentaro Mori, Hiroshi Kagami, Hidemichi Ito, Hidetaka Onodera, Yasuyuki Kaga, Haruki Ohtsubo, Kentaro Tatsuno, Noriko Usuki, Satoshi Takaishi, Yoshihisa Yamano, the K-NET Registry Investigators

## Abstract

**Background:** The benefits of endovascular thrombectomy (EVT) for distal medium vessel occlusions (DMVOs) are not well established. This study aimed to evaluate whether EVT is superior to intravenous tissue plasminogen activator (IV tPA) alone in DMVOs.

**Methods:** This study analyzed data from the K-NET Registry, a prospective, multicenter, observational registry of acute ischemic stroke patients treated with EVT or IV tPA. The study evaluated patients with acute DMVOs who were treated with EVT and/or IV tPA. DMVOs was defined as occlusions in M2-M3 segment of the middle cerebral artery, anterior cerebral artery, or posterior cerebral artery. The analysis included primary DMVOs and excluded secondary DMVOs, such as distal embolism after recanalization of proximal vessel occlusion. Propensity score-matched analysis was conducted to compare the outcomes between EVT and IV tPA alone. A good outcome was defined as a modified Rankin Scale score of 0-2 or no worsening at 90 days. An excellent outcome was defined as an mRS score of 0-1.

**Results:** The study included 1148 patients with DMVOs, of whom 816 were treated with EVT and 332 were IV tPA alone. Before propensity score matching, the incidence of good and excellent outcomes was significantly lower in EVT group (good outcomes: EVT 50.3% vs. IV tPA 68.0%, p<0.01; excellent outcomes: 39.8% vs. 59.8%, p<0.001). After propensity score matching, there were no significant differences between EVT and IV tPA groups in good outcomes (EVT 57.8% vs. IV tPA 61.3%, p=0.51), excellent outcomes (46.6% vs. 55.0%, p=0.17), all cerebral hemorrhage (11.6% vs. 12.7%, p=0.74), and symptomatic hemorrhage (2.9% vs. 0.6%, p=0.13). Subarachnoid hemorrhage was more frequent in EVT group (14.5% vs. IV tPA 0%).

**Conclusions:** The benefits of EVT for acute DMVOs were similar to IV-tPA alone. Randomized multicenter trials are warranted to establish the superiority of EVT over IV-tPA alone for DMVOs.

## Introduction

Endovascular thrombectomy (EVT) has become the gold standard treatment for stroke patients with acute large vessel occlusion. However, there is limited evidence for the benefits of EVT for more distal, medium vessel occlusions (DMVOs). The first five landmark EVT trials of internal carotid artery (ICA) or middle cerebral artery (MCA) occlusion stroke included only 7% of patients with M2 segment occlusion, and patients with M3 occlusion were not included. ^1^ Two randomized controlled trials investigating EVT for late-onset anterior circulation stroke did not include M2 or M3 occlusions. ^2,3^ Moreover, the efficacy of EVT versus medical treatment in anterior cerebral artery (ACA) and posterior cerebral artery (PCA) occlusion, even in A1 and P1 occlusions, has not been evaluated in these randomized controlled trials. According to guidelines for acute ischemic stroke, EVT is strongly recommended for ICA or MCA M1 occlusion, while EVT for M2 or M3 occlusion is given a weak recommendation. No recommendation is given for ACA and PCA occlusion due to the lack of evidence. ^4,5^

Intravenous tissue-type plasminogen activator (IV tPA) is one of the standard treatments for DMVOs. The incidence of recanalization after IV tPA for DMVOs was expected to be higher than that for large vessel occlusion. However, the recanalization rate of IV tPA for DMVOs remains inadequate. A previous meta-analysis reported a complete recanalization rate of 38% and a partial or complete recanalization rate of 52% for M2-M3 occlusion. ^6^ The INTERRSeCT study reported recanalization incidences of 43% after IV tPA for M3, ACA, or PCA occlusions. ^7^ Several studies have reported high recanalization rates with EVT for DMVOs, but it remains unclear whether EVT provides higher recanalization rates and superior outcomes compared to IV tPA alone. There are some studies that compared the outcome of EVT and medical management for DMVOs, however, most of the previous studies included patients treated without IV tPA as a medical management arm. Since IV tPA has been shown to lead to better clinical outcomes than treatment without IV tPA, it is important to differentiate medical management with and without IV tPA when comparing it with EVT.

This study analyzed a prospective stroke registry of patients who underwent EVT or IV tPA for acute DMVOs. We aimed to evaluate whether the outcomes of acute ischemic stroke patients treated with EVT for DMVOs were superior to those treated with IV tPA alone.

## Methods

### Study Population

The study population consisted of patients enrolled in the Kanagawa Intravenous and Endovascular Treatment of Acute Ischemic Stroke Registry (K-NET Registry), which is a prospective, multicenter, observational study conducted in 40 stroke centers in Kanagawa Prefecture, Japan. This registry aimed to investigate patients with acute ischemic stroke who received intravenous tPA therapy and/or endovascular therapy. The detailed protocol and primary results of the K-NET registry have been published previously. ^8^ The K-NET registry included patients who received intravenous tPA therapy for acute ischemic stroke and/or intended to undergo endovascular therapy for large vessel occlusion. The K-NET Registry collected basic information, such as patient background, imaging findings, procedure details, time of treatment, perioperative complications, and mRS at 90 days after stroke onset. Intracranial hemorrhage was classified by the Safe Implementation of Thrombolysis in Stroke-monitoring Study criteria. ^9^ The data underwent standardized quality checks to control for consistency, plausibility, and completeness. In the K-NET Registry, there were no missing data and no cases with loss to follow-up at 90 days. The data supporting the findings of this study are available from the corresponding author upon reasonable request.

The present study is based on the patients’ data in the K-NET registry between January 2018 and December 2021. The inclusion criteria for the present analysis were as follows: (1) primary DMVOs and (2) treated with EVT and/or IV tPA. DMVOs was defined as MCA M2-M3, ACA A1-A3, and PCA P1-P3 occlusion. The occlusion site was identified using MRA or CTA in IV tPA alone cases and digital subtraction angiography in EVT cases. The analysis included only primary DMVOs and excluded secondary DMVOs, such as distal embolism occurring after recanalization of proximal large vessel occlusion.

### Ethics Approval

The study protocol of the K-NET registry was approved by the Ethics Committee of St. Marianna University School of Medicine (approval no. 3757). In accordance with local regulations, further approval was obtained from relevant local institutional ethics committees or institutional review boards. Written informed consent was obtained from each patient or their surrogate prior to their inclusion in the registry. The necessary information for registration was obtained as part of routine clinical practice.

### Imaging and Endovascular Treatment

Upon admission, patients underwent diagnostic imaging using either CT or MRI. The specific imaging modality employed to assess thrombectomy-capable patients (MRI, CTA, or CTP) was determined at each participating institution. Patients with indications for IV t-PA were treated with alteplase at a dose of 0.6 mg per kilogram of body weight according to Japanese guidelines at the discretion of the treating physician. The choice of EVT technique, including stent retriever, aspiration, percutaneous transluminal angioplasty, and intra-arterial thrombolysis, was determined by neurointerventionalists at each institution. There were no restrictions on the selection of EVT devices. Recanalization grade was measured using the modified Thrombolysis in Cerebral Infarction (mTICI) scale evaluated by experienced neurologists, neurosurgeons, or neuroradiologists in each institution.

### Outcome

The primary outcome was good outcome defined as modified Rankin Scale (mRS) 0-2 at 90 days after the stroke onset. As this study also included patients with pre-stroke mRS 3-5, no decrease in mRS score at 90 days was also defined as a good outcome, but only in patients with pre-stroke mRS 3-5, in concordance with the definitions used in previous reports. ^8,10^ The safety outcome was measured by all cerebral hemorrhage, symptomatic hemorrhage, and subarachnoid hemorrhage. The secondary efficacy outcome was excellent outcome defined as the mRS 0-1, excluding patients with pre-mRS of more than two for this evaluation.

### Statistical analysis

The baseline characteristics of the patients were analyzed using standard statistical methods. Continuous variables were described using mean and standard deviation (SD), while ordinal variables were presented as median with interquartile range (IQR). Categorical variables were expressed as proportions. Univariate analysis was performed using the χ2 test or Fisher exact test, as appropriate.

To reduce the possibility of selection bias, propensity score matching (PSM) was performed to adjust for covariates of baseline variables. PSM was performed based on 1:1 pair matching without replacement using the nearest-neighbor matching algorithm. Covariates for PSM were age, sex, pre-stroke mRS, NIHSS, stroke cause, occlusion site, DWI ASPECTS, and time from symptom recognition to hospital arrival. We repeated the same PSM procedure to evaluate excellent outcomes after excluding patients with a pre-mRS score greater than two. The caliper width was set to the highest value between 0.2 and 0.01, ensuring that all selected baseline covariates had an absolute standardized difference of ≤ 0.1. If the standardized difference was greater than 0.1 for each caliper widths between 0.2 and 0.01, the caliper width was set to the value that yielded the minimum standardized difference. As a result, caliper widths of 0.11 and 0.06 were selected for PSM in all patients and evaluation of excellent outcomes excluding pre-mRS score greater than two, respectively.

EVT and IV tPA alone groups were compared before and after PSM using binary logistic regression to assess the association between treatment and outcome. Subgroup analysis was performed according to occlusion site (MCA M2, MCA M3, ACA or PCA occlusion), stroke severity (NIHSS ≥10 or ≤9), EVT with/without IV tPA, thrombectomy methods (stent retriever or aspiration only) and symptom recognition to door time <4.5 hours. For subgroup analysis, we divided each subgroup from a matching cohort and performed multivariate analysis to evaluate whether EVT or IV tPA alone treatment was associated with outcomes. Odds ratios, confidence intervals, and corresponding p-values were derived, with a p-value < 0.05 considered significant. Statistical analyses were conducted using SAS version 9.4 (SAS institute, Cary, NC, USA) and Stata version 17.0 (Stata Corporation, College Station, TX, USA). PSM was conducted using SAS version 9.4 (SAS institute, Cary, NC, USA).

## Results

Between January 2018 and December 2021, 3954 patients who were treated with EVT or IV tPA were included in the K-NET registry. Among them, 2806 patients without distal vessel occlusion were excluded. Finally, 1148 patients with DMVOs were included in this analysis. Of these, 889 patients with pre-mRS 0 or 1 were included for evaluating excellent outcome. The flowchart is shown in **Figure 1**.

**Figure 1.**
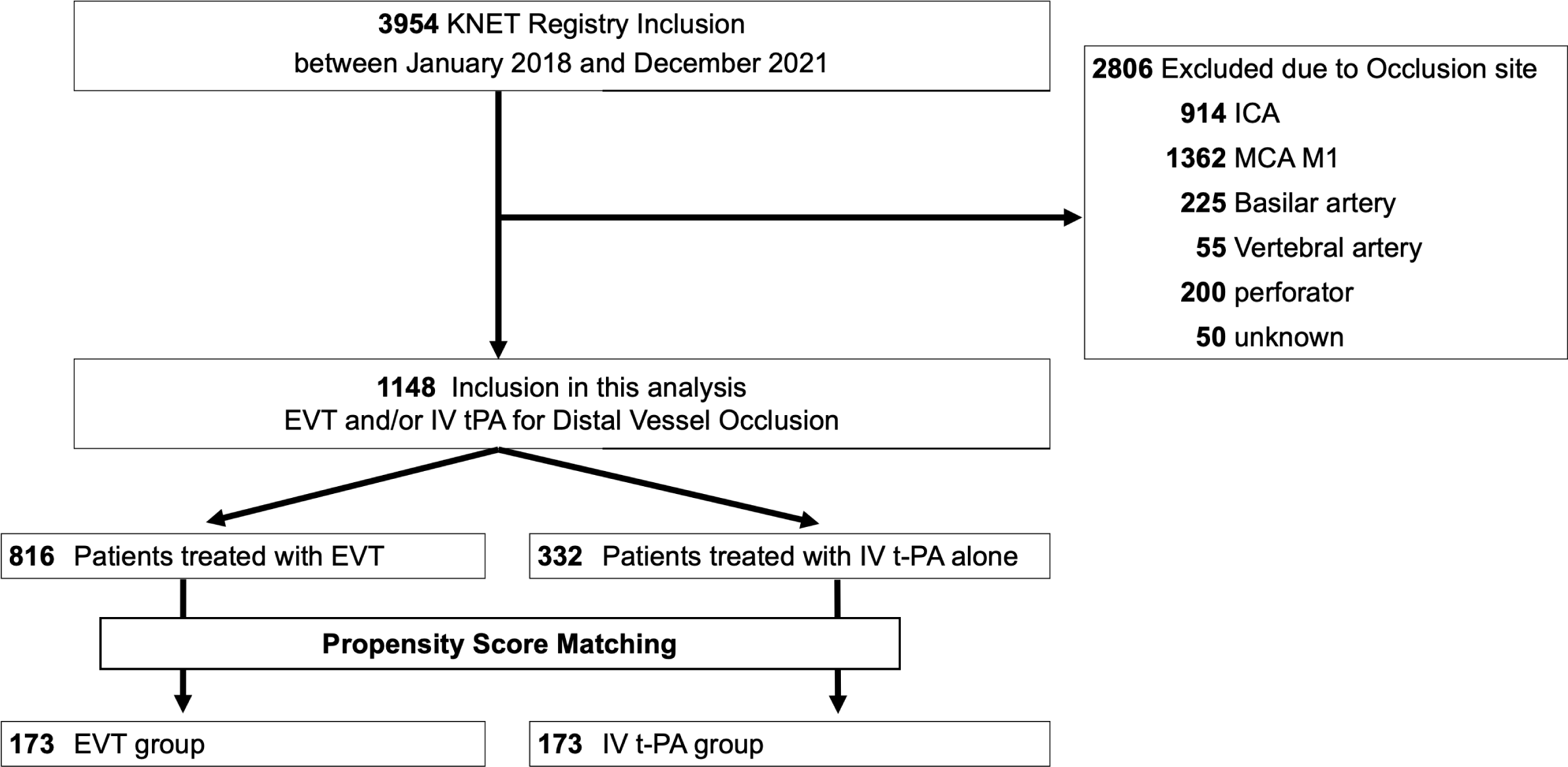
Study flowchart The inclusion criteria for the K-NET registry encompassed patients who received intravenous tPA therapy for acute ischemic stroke and/or intended to undergo endovascular therapy for large vessel occlusion. There were no specific exclusion criteria. EVT indicates Endovascular thrombectomy; IV tPA, Intravenous tissue-type plasminogen activator; ICA, internal carotid artery; MCA, middle cerebral artery.

### Baseline Characteristics

Of 1148 patients with DMVOs included in the current analysis, 816 patients received EVT, and 332 patients received IV tPA alone. Baseline characteristics in each group are shown in **Table 1**. The EVT group had a higher prevalence of diabetes (EVT 18.0 % vs. IV tPA 12.7%; P=0.02), atrial fibrillation (49.7% vs. 38.9%; P<0.01), higher NIHSS (median [IQR], 14.5 [9-22] vs. 9 [5-15]; P <0.01) and longer onset-door time (68 minutes [45.5-116.5] vs. 58.5 minutes [42-92.5]; P <0.01). The EVT group had a significantly higher frequency of M2 occlusion (78.7% vs. 34.6%; P<0.01), while the IV tPA group had a higher frequency of M3 occlusion (8.8% vs. 44.3%; P<0.01).

**Table 1.**
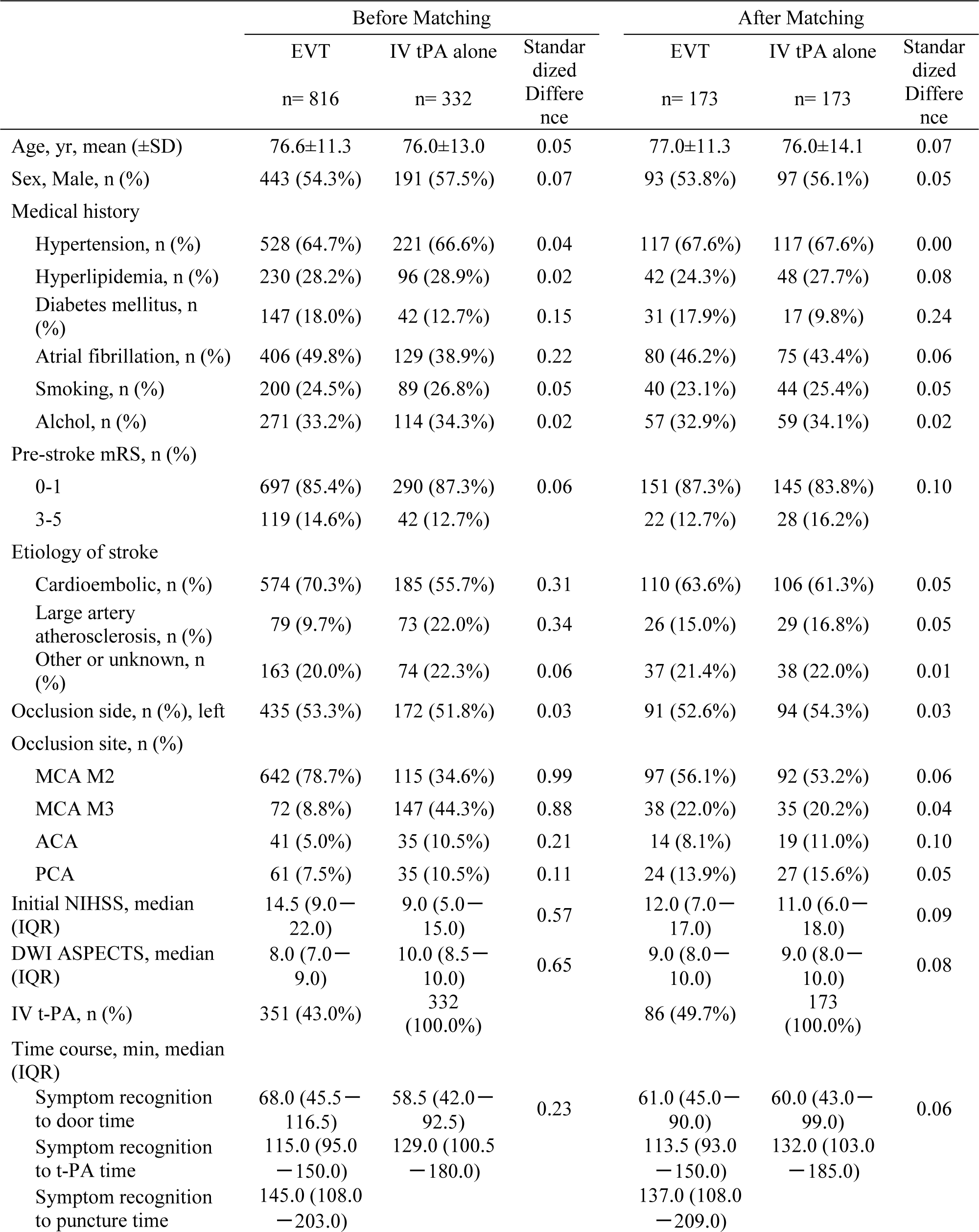
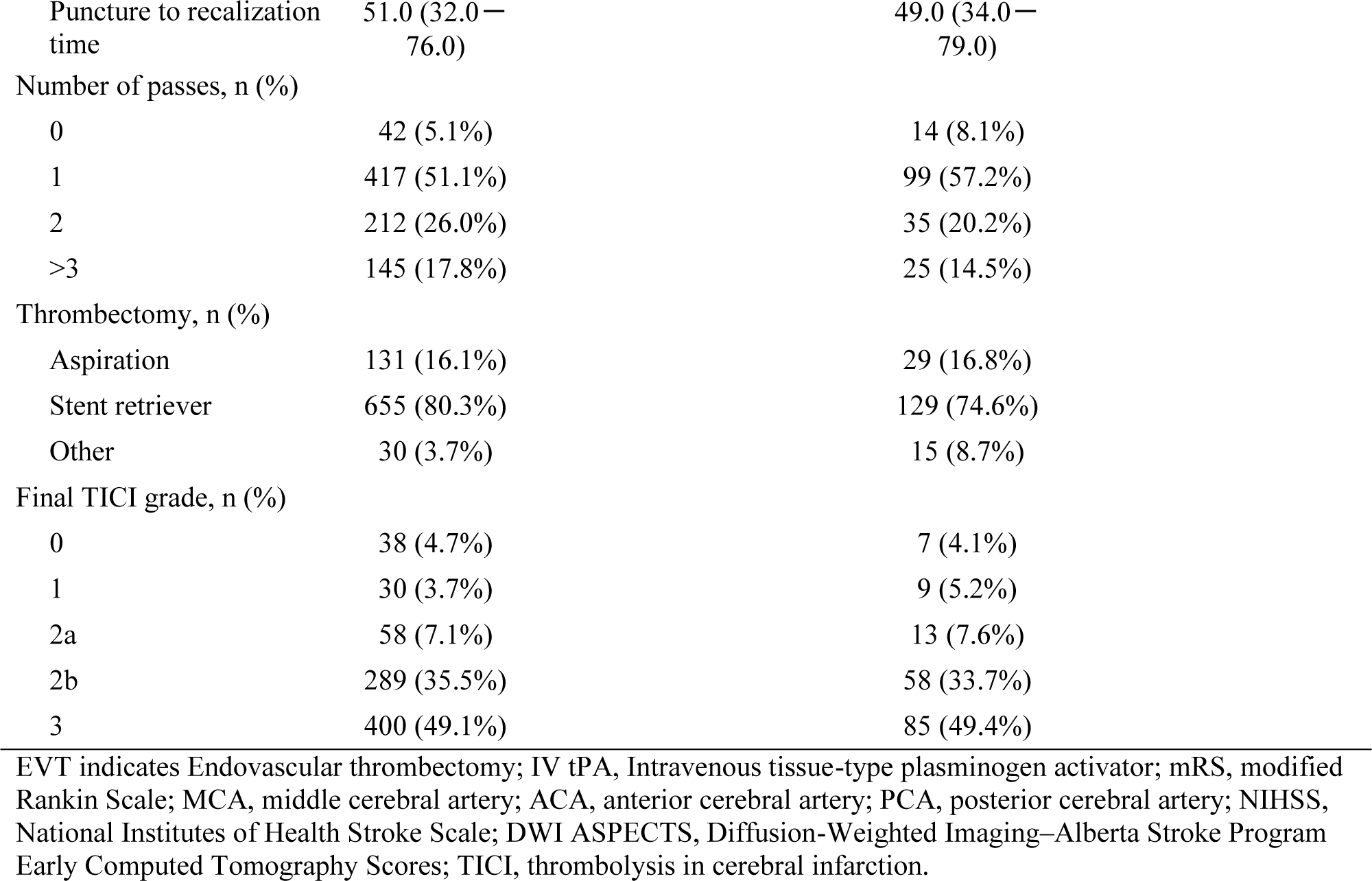
Baseline characteristics between endovascular thrombectomy group and IV t-PA alone group before and after propensity score matching.

In the EVT group, the median time from symptom recognition to groin puncture was 145 minutes (IQR, 108-203 minutes). 351 (43.0%) patients were treated by EVT with IV tPA and 465 (57.0%) were EVT alone. Final TICI 2b or more was achieved in 689 (84.5%) and TICI 3 was 400 (49.1%) patients. A stent retriever with/without an aspiration catheter was used in 655 (80.3%), aspiration only in 131 (16.1%), and 30 (3.7%) patients were treated with other techniques including intra-arterial thrombolysis. In patients treated with stent retriever or aspiration, the number of passes was once in 417 (51.1%), twice in 212 (26.0%), and more than three in 145 (17.8%) patients. The median time from puncture to recanalization was 51 minutes (IQR, 32-76 minutes)

### Clinical and safety outcomes before propensity score matching

Before PSM, the incidence of good clinical outcomes was significantly lower in the EVT group (EVT 50.3% vs. IV tPA 68.0%; p<0.01, **Figure 2, Table2**). There was a trend of increasing all cerebral hemorrhage (14.8% vs. 11.1%; p=0.10). No significant difference was found in the frequency of symptomatic hemorrhage (3.1% vs. 2.4%; p=0.55). Subarachnoid hemorrhage was significantly more frequent in the EVT group (17.8% vs. 0%). After excluding patients with pre-stroke mRS of more than two, excellent outcomes were significantly lower in the EVT group (39.8% vs. 59.8%; p<0.001).

**Figure 2.**
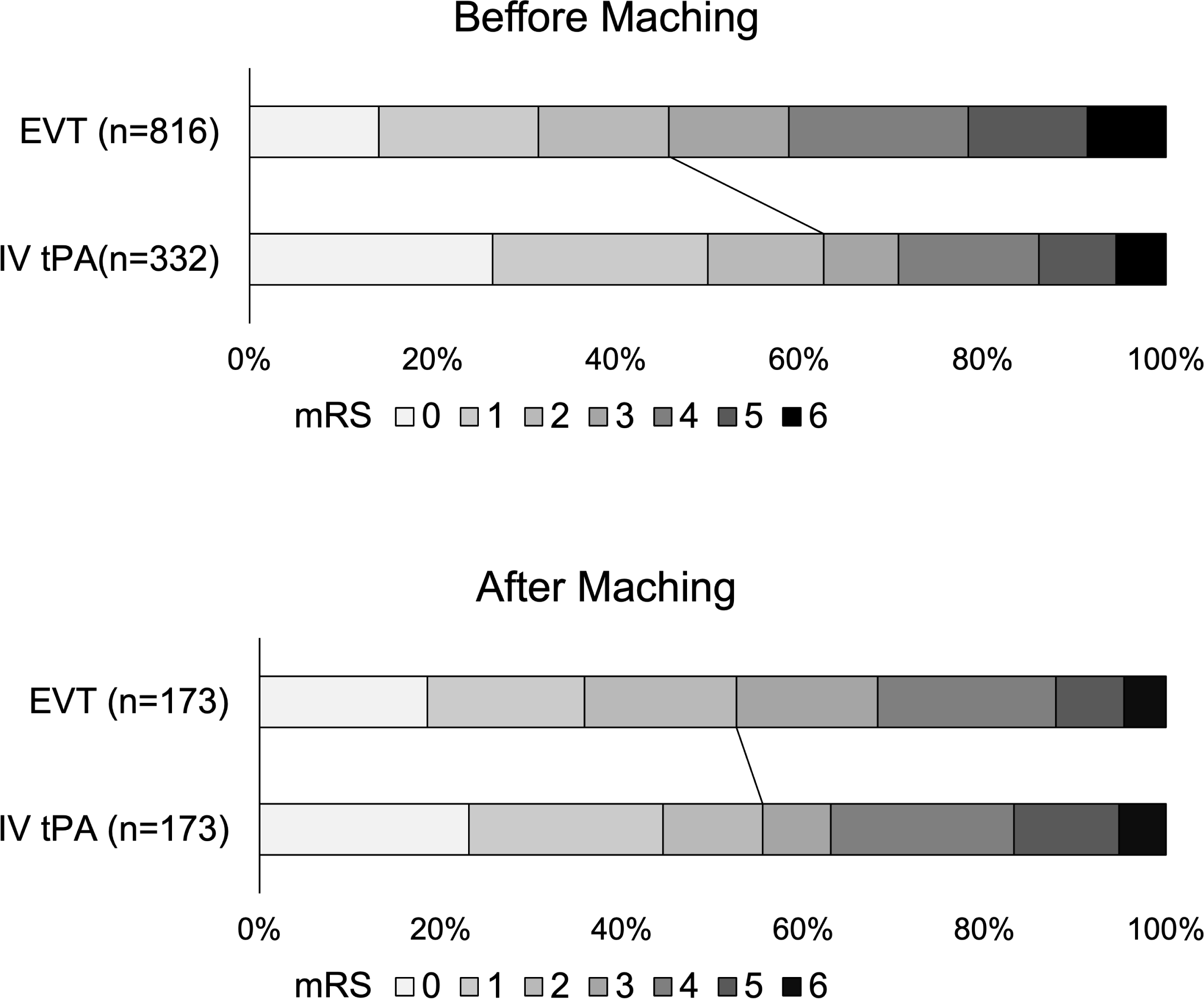
Distribution of modified Rankin Scale Score at 90 days before and after propensity score matching EVT indicates Endovascular thrombectomy; IV tPA, Intravenous tissue-type plasminogen activator; mRS, modified Rankin Scale.

**Table 2.** Efficacy and Safety Outcome between EVT and IV tPA before and after Propensity Score Matching.

|  | Before Matching |  |  |  | After Matching |  |  |  |
| --- | --- | --- | --- | --- | --- | --- | --- | --- |
|  | IV tPA | EVT | OR (95% CI) | P value | IV tPA | EVT | OR (95% CI) | P value |
| All Patients |  |  |  |  |  |  |  |  |
| n | 332 | 816 |  |  | 173 | 173 |  |  |
| Good outcome | 226 (68.0%) | 410 (50.3%) | 0.474 (0.362-0.620) | <0.01 | 106 (61.3%) | 100 (57.8%) | 0.866 (0.562-1.331) | 0.51 |
| All cerebral hemorrhage | 37 (11.1%) | 121 (14.8%) | 1.388 (0.937-2.055) | 0.10 | 22 (12.7%) | 20 (11.6%) | 0.897 (0.470-1.712) | 0.74 |
| Symptomatic hemorrhage | 8 (2.4%) | 25 (3.1%) | 1.280 (0.571-2.867) | 0.55 | 1 (0.6%) | 5 (2.9%) | 5.119 (0.592-44.278) | 0.13 |
| Subarachnoid Hemorrhage | 0 (0%) | 145 (17.8%) | N/A | N/A | 0 (0%) | 25 (14.5%) | N/A | N/A |
| Patients with Pre-mRS 0-1 |  |  |  |  |  |  |  |  |
| n | 271 | 618 |  |  | 131 | 131 |  |  |
| Excellent outcome | 162 (59.8%) | 246 (39.8%) | 0.445 (0.332-0.595) | <0.01 | 72 (55.0%) | 61 (46.6%) | 0.714 (0.439-1.161) | 0.17 |
| All cerebral hemorrhage | 28 (10.3%) | 81 (13.1%) | 1.309 (0.830-2.064) | 0.25 | 14 (10.7%) | 14 (10.7%) | 1.000 (0.457-2.190) | 1.00 |
| Symptomatic hemorrhage | 6 (2.2%) | 19 (3.1%) | 1.400 (0.553-3.546) | 0.48 | 1 (0.8%) | 2 (1.5%) | 2.015 (0.181-22.503) | 0.56 |
| Subarachnoid Hemorrhage | 0 (0%) | 107 (17.3%) | N/A | N/A | 0 (0%) | 22 (16.8%) | N/A | N/A |

### Clinical and safety outcomes after propensity score matching

After PSM, 173 matched pairs were identified. Baseline characteristics between the groups achieved good balance (**Table 1**). There were no differences in good clinical outcome (EVT 57.8% vs. IV tPA 61.3%; p=0.51), all cerebral hemorrhage (11.6% vs. 12.7%; p=0.74), and symptomatic hemorrhage (2.9% vs. 0.6%; p=0.13) between EVT and IV tPA group (**Figure2, Table2**). Subarachnoid hemorrhage was significantly frequent in the EVT group (EVT 14.5%, IV tPA 0%).

In PSM excluding patients with pre-mRS more than two to evaluate excellent outcomes, 131 matched pairs were identified. Baseline characteristics between the groups achieved good balance (**Supplemental Table I**). There were no significant differences in excellent outcomes between EVT and IV tPA groups (EVT 46.6%, IV tPA 55.0%; p=0.17).

### Subgroup Analysis

Subgroup analysis after PSM was shown in **Figure 3, Supplemental Table 2 and 3**. Aspiration-only treatment in the EVT group had a significantly higher rate of good outcomes compared to IV t-PA group (Odds ratio (95% CI) 9.555 (2.066-44.193); p<0.01). In other subgroup, there were no significant differences between the two-treatment cohorts for good outcomes, all cerebral hemorrhage, and symptomatic hemorrhage. Subarachnoid hemorrhage was significantly higher in all subgroups except for ACA occlusion, PCA occlusion, and aspiration group.

**Figure 3.**
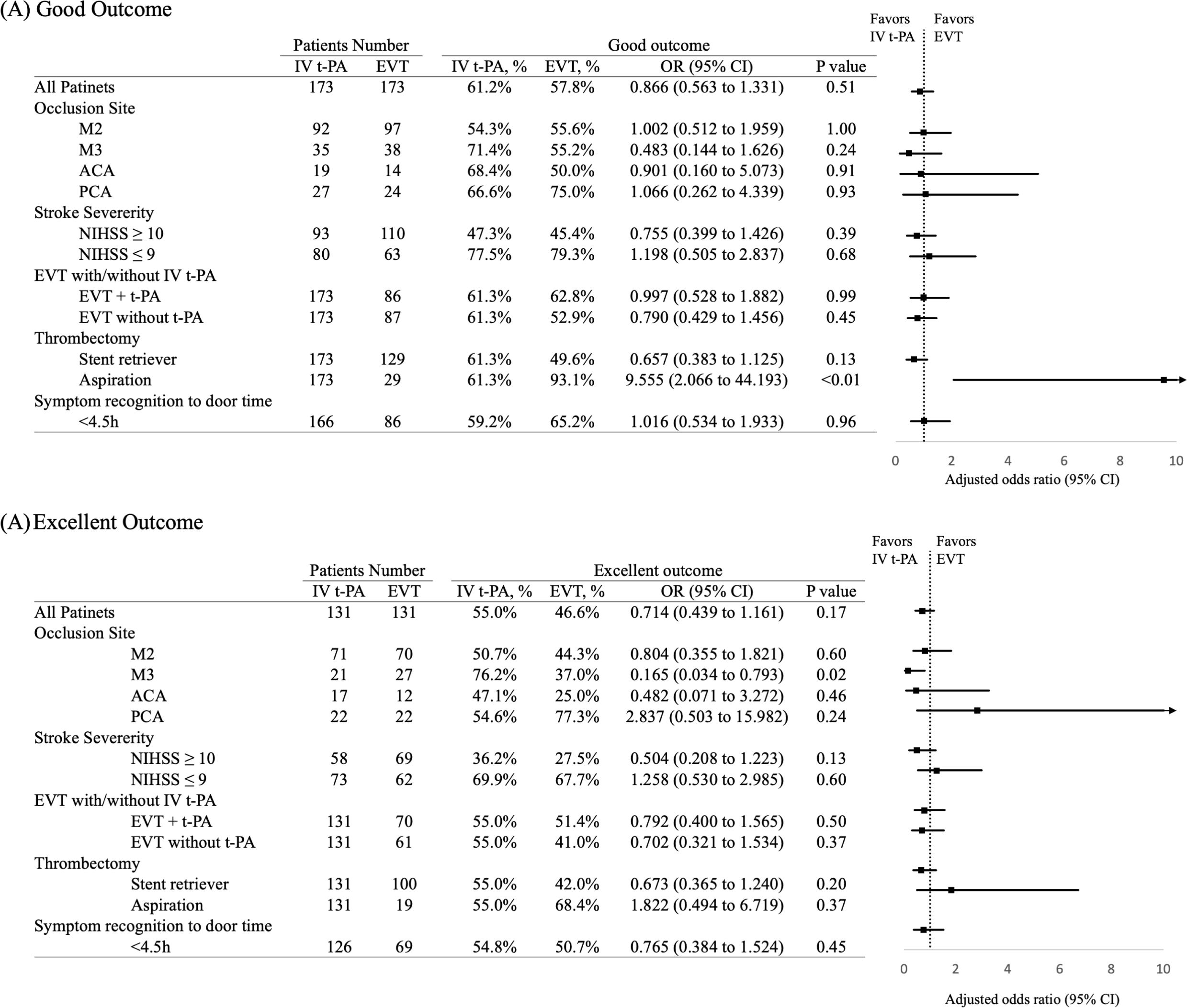
Subgroup analyses after propensity score matching for clinical outcome and hemorrhage The forest plot demonstrates the odds ratio of good clinical outcome (A) and excellent outcome (B) after propensity score matching. Good outcome was defined as mRS 0-2 at 90 days, or no decrease in mRS score at 90 days in patients with pre-stroke mRS 3-5. Excellent outcome was defined as the mRS 0-1, excluding patients with pre-mRS of more than. Arrows indicate that the limits of the confidence interval are not fully shown. EVT indicates Endovascular thrombectomy; IV tPA, Intravenous tissue-type plasminogen activator; OR, odds ratio; MCA, middle cerebral artery; ACA, anterior cerebral artery; PCA, posterior cerebral artery; NIHSS, National Institutes of Health Stroke Scale.

After PSM excluding patients with pre-mRS more than two, EVT group had a significantly lower rate of excellent outcomes than the IV tPA group in M3 occlusion group (Odds ratio (95% CI) 0.165 (0.034-0.793); p=0.02). In other subgroups, there were no significant differences in excellent outcome between the two treatment cohorts.

## Discussion

In this PSM analysis, we compared the outcomes of EVT versus IV tPA alone for acute primary DMVOs. Our findings demonstrated no significant differences in efficacy between EVT and IV tPA alone. Subgroup analysis further supported these results, indicating no significant differences in efficacy within most of the subgroups. Though aspiration-only treatment had a significantly better outcome than IV tPA alone and IV tPA alone had a significantly better outcome in the subgroup of M3 occlusion, it is important to note that the number of patients in these subgroups was limited, with only 29 patients for the aspiration-only group and 27 for the EVT group in M3 occlusion after PSM. Regarding safety, there were no significant differences in symptomatic hemorrhage, while subarachnoid hemorrhage was significantly more frequent in the EVT group. These results suggest that IV tPA remains a first-line treatment for primary distal occlusions. When considering the addition of EVT to IV tPA, careful evaluation is necessary as the effectiveness of EVT with IV tPA is similar to that of IV tPA alone. On the other hand, since the safety of EVT without IV tPA was similar to IV tPA alone, EVT is a feasible treatment for patients with contraindications to IV tPA.

In the current study, we defined DMVOs as MCA M2, M3, ACA, and PCA occlusion, which included A1 in ACA and P1 in PCA occlusions. The Distal Thrombectomy Summit Group has recommended considering two anatomical features, vessel size and distance/tortuosity, for the classification of DMVOs. ^11^ There is wide agreement on the inclusion of distal, medium artery categories, including the MCA M3 to M4, ACA A2 to A5, and PCA P2 to P5 segments. However, the categorization of MCA M2, ACA A1, and PCA P1 occlusions has been variable. Some previous studies that investigated EVT for DMVOs included these vessel occlusions as DMVOs, in contrast, other studies for DMVOs excluded these vessels. ^12^ Although the definition of DMVOs remains debatable, there is currently insufficient strong evidence supporting EVT for M2, A1, and P1 occlusions. Thus, our study included these vessel occlusions as DMVOs. To address this issue, we also conducted a subgroup analysis based on vessel occlusion, distinguishing between M2, M3, ACA, and PCA occlusions.

There have been several studies comparing EVT with medical management for DMVOs. While the subgroup analysis in HERMES collaboration showed the direction of effect favored EVT in M2 occlusion without statistical significance, ^1^ patient-level data indicated that EVT favored good clinical outcomes at 90 days for patients with proximal M2 occlusions eligible for EVT trial protocols. ^13^ It is worth noting that most of these randomized control trials excluded patients with M2 occlusions, resulting in a small sample size of only 67 patients in the EVT arm. A multicenter retrospective cohort study assessing patients with acute isolated M2 segment occlusions showed that EVT-treated patients had three times the odds of good outcomes compared to those who received medical management. This treatment effect remained significant even after adjustment for age, NIHSS score, ASPECTS, IV tPA treatment, and time from last known normal to arrival at the emergency department. ^14^ In contrast, some previous studies revealed no superiority of EVT compared to medical management. A multicenter cohort study that pooled data from patients with primary anterior circulation DMVOs, defined as any segment of the ACA or distal MCA, showed no significant differences in 3-month functional independence between the EVT and medical management groups. ^15^ Regarding ACA occlusions, a previous registry study for ACA A2 or A3 occlusion reported similar favorable functional outcomes and mortality rates between EVT and medical management groups after PSM. ^16^ For PCA occlusions, a multicenter case-control study analyzing patients with primary distal occlusion of the P2 or P3 segment showed that the mRS score distributions at the 90-day follow-up did not differ significantly between the EVT and medical treatment cohorts. Significant treatment effects of mechanical thrombectomy were observed in a subgroup of patients with higher NIHSS scores of 10 or higher on admission and who were not eligible for IVT. ^17^ Another observational study of patients with proximal PCA occlusion showed that EVT was not associated with good or excellent functional outcomes compared to the best medical treatment alone. In addition, this study showed EVT was associated with higher rates of symptomatic intracranial hemorrhage and early neurological deterioration. ^18^ Similarly, a case-control study evaluating patients with primary PCA occlusion also showed that EVT was associated with similar odds of disability by ordinal mRS and similar rates of functional independence, despite a higher rate of symptomatic intracranial hemorrhage and mortality. However, this study found a higher likelihood of excellent outcomes, early NIHSS improvement, and complete vision recovery compared to medical management.^19^

Therefore, previous studies on DMVOs have produced inconsistent results, with some showing the superiority of EVT and others not demonstrating superiority. A meta-analysis that included 14 observational and two randomized controlled studies of distal medium vessel occlusions found that EVT achieved significantly better odds of functional independence compared to the best medical treatment. ^12^ However, there was a high degree of heterogeneity between studies that may confound funnel plot symmetry. Additionally, there were no significant differences in excellent functional outcomes. Consequently, these previous reports provide insufficient evidence regarding the benefit of EVT over medical management for DMVOs. Our study also found no clear evidence for the superiority of EVT over IV tPA alone.

Strengths of our study include our study design using PSM from the large number of patients with DMVOs to reduce the possibility of selection bias. Another strength is that we compared EVT and IV tPA groups, excluding patients treated without IV tPA from the medical management group. In previous studies that compared EVT and medical management for DMVOs, approximately 25% (0-60%) of patients in the medical management group were treated without IV tPA. There are several limitations in our study. Firstly, it is a retrospective analysis of a single-arm prospective registry. Potential sources of bias, such as selection bias and measurement bias, were identified in our retrospective analysis of the registry study. To address these potential biases, we utilized PSM to adjust for confounding variables. In addition, subgroup analysis was performed to assess for variability and consistency of results within each subgroup. Moreover, a standardized protocol for data collection was implemented to minimize measurement bias. Another bias is the bias in EVT method including stent retriever and aspiration only, which was determined by the neurointerventionalists at each institution. To address this potential bias, we performed subgroup analysis according to EVT methods. The second limitation is the lack of data regarding the specific segment of M2 occlusion (proximal or distal side). Previous studies have shown that patients with proximal M2 segment MCA occlusions eligible for EVT trial protocols benefited from EVT. ^13^ Similarly, there is a lack of data regarding the segment of the anterior cerebral artery (A1, A2, or A3) and posterior cerebral artery (P1, P2, or P3) occlusions. Lastly, our study included patients with pre-stroke mRS scores of 2 or more, and we defined a good clinical outcome as an mRS score of 0-2 or no decrease in the mRS score at 90 days after stroke. The common definition of a good clinical outcome is mRS score of 0-2, as many previous studies excluded patients with pre-stroke mRS scores of 2 or more. To address this limitation, we also evaluated excellent outcomes defined as mRS 0-1 at 90 days, excluding patients with pre-stroke mRS scores of 2 or more, and found no significant difference between the EVT and IV tPA groups.

In conclusion, the outcome of EVT for acute primary DMVOs was similar to IV-tPA alone. The extent to which EVT brings benefits may differ between proximal large vessel occlusion and DMVOs. Randomized multicenter trials are warranted to establish the superiority of EVT over IV-tPA alone for DMVOs.

## Data Availability

The data supporting the findings of this study are available from the corresponding author upon reasonable request.

## Acknowledgments

We would like to thank the K-NET Registry investigators. We are grateful to Ms. Tomomi Shibuya and Mr. Satoshi Muta (Department of Practical Management of Medical Information, St. Marianna University School of Medicine) for collecting and checking the data.

